## Supplemental tables for "Oxygen provision to severely ill COVID-19 patients at the peak of the 2020 pandemic in a Swedish district hospital": Supplementary tables_Hvarfner.pdf

**Supplementary table 1** Patient characteristics and outcomes

| | Patients admitted to the wards who never received oxygen treatment<br>% (n/N), unless otherwise stated | Patients initially admitted to the wards and who were later transferred to ICU<br>% (n/N), unless otherwise stated |
|---|---|---|
| **Age (years), median (IQR)** | 60 (52-73) | 61 (56-70) |
| **Female** | 52% (25/48) | 22% (4/18) |
| **Diagnosis of COVID-19 confirmed by PCR** | 81% (39/48) | 100% (18/18) |
| **BMI, ≥30** | 23% (7/30) | 36% (4/11) |
| **CACI ≥4** | 35% (17/48) | 17% (3/18) |
| **No-ICU-decision documented , n (%)** | 31% (14/48) | 11% (2/18) |
| *Red NEWS-2 (1) parameter on first measurements of vital signs* | | |
| **SpO2** (≤91%) | 2.1% (1/48) | 56% (10/18) |
| **Respiratory rate** (≤8 or ≥25 breaths/min**)** | 24% (11/45) | 44% (8/18) |
| **Heart rate** (≤40 or ≥131 beats/min) | 2.1% (1/48) | 0% (0/18) |
| **Systolic blood pressure** (≤90 or ≥220 mmHg) | 2.1% (1/48) | 0% (0/18) |
| **Consciousness** (Non-alert) | 6.4% (3/47) | 0% (0/18) |
| **Temperature** (≤35.0 or ≥39.1°C) | 8.3% (4/48) | 44% (8/18) |
| *Treatments during hospital-stay* | | |
| **Antibiotics** | 56% (27/48) | 100% (18/18) |
| **Chloroquine** | 6.2% (3/48) | 28% (5/18) |
| **Anticoagulants** | 40% (19/48) | 94% (17/18) |
| *Outcomes* | | |

| | | |
|---|---|---|
| **Length of stay (days), median (IQR)** | 1.9 (1.1-3.1) | 24 (9.5-31) |
| **Transfer to another department** | 4% (2/48) | 50% (9/18) |
| **Dead in-hospital** | 0% (0/48) | 22% (4/18) |
| **Dead at 60 days** | 0% (0/48) | 22% (4/18) |

*Abbreviations: PCR: polymerase chain reaction, BMI: body mass index, CACI: Charlson´s age adjusted comorbidity score, ICU: intensive care unit, SpO2: peripheral oxygen saturation

** For two patients the initial no-ICU decisions were changed after time and the patients were transferred to ICU.

[1] Royal College of Physicians. National Early Warning Score (NEWS) 2. Standardising the assessment of acute-illness severity in the NHS. Updated report of a working party. [Internet]. London: RCP; 2017 [cited 2021 Jan 6]. Available from: https://www.rcplondon.ac.uk/projects/outputs/national-early-warning-score-news-2

**Supplementary table 2** Clinical progression score for all admitted patients[2]

| Descriptor | Score | All admitted patients (n=206) | All admitted patients aged <70 (n=120) | All admitted patients with no-ICU-decision (n=77) |
|---|---|---|---|---|
| Hospitalized without oxygen | 4 | 23% (48/206) | 29% (35/120) | 19% (15/77) |
| Hospitalized with oxygen nasal spongs or mask | 5 | 43% (88/206) | 49% (59/120) | 27% (21/77) |
| Hospitalised; oxygen by NIV or high flow | 6 | 1% (2/206) | 2% (2/120) | 0% (0/77) |
| Intubation and mechanical ventilator | 7 to 9 | 10% (20/206) | 12% (14/120) | 1% (1/77) |
| Dead after 60 days | 10 | 23% (48/206) | 8.3% (10/120) | 52% (40/77) |

[2] Clinical progression score, adapted from Marshall JC, Murthy S, Diaz J, Adhikari NK, Angus DC, Arabi YM, et al. A minimal common outcome measure set for COVID-19 clinical research. Lancet Infect Dis. 2020 Aug 1;20(8):e192–7.

*Initial no-ICU decision changed after time and the patient transferred to ICU.

**Supplementary table 3** Oxygen provision in the wards to patients that were later transferred to ICU

| | **Patients initially admitted to the wards that later transferred to ICU (N=18)** |
|---|---|

| | |
|---|---|
| **Days on oxygen treatment before transfer to ICU, median (IQR)** | 1.8 (0.32-5.0) |
| **Oxygen flow to patients during oxygen therapy (l/min)** | |
| - Mean (SD) | 7.5 (3.4) |
| - Median (IQR) | 7.5 (4.5-9.3) |
| **Oxygen flow to patients during time in the ward (l/min)** | |
| - Mean (SD) | 6.7 (4.0) |
| - Median (IQR) | 7.0 (3.3-8.5) |
| **Total volume of oxygen provided per patient admission (l)** | |
| - Mean (SD) | 34,000 (51,000) |
| - Median (IQR) | 13,000 (4,100-59,000) |
